# An LLM-Based Auditing Framework for Targeted Quality Assurance in Cancer Registries

**DOI:** 10.64898/2026.09.22.26363595

**Authors:** Maryam Seifaddini, Mohammad Beheshti, Jeffrey Steffens, Deborah Carnagey, Magda Esebua, Mark Wakefield, Iris Zachary

## Abstract

Cancer registries are essential for population-level surveillance, epidemiologic research, and public health planning, but their accuracy depends on human review of key variables documented in unstructured clinical text. This is time-consuming, and current quality assurance (QA) typically relies on reviewing fewer than 10% of records at random, which can miss facility- or time-specific errors.

We developed a large language model (LLM)-based framework for targeted cancer registry QA. The framework uses LLMs to extract biomarker values from clinical text, compares them with registry entries, and flags disagreements for expert review. Six open-source LLMs were benchmarked locally in a privacy-preserving environment, and the best model per site and report type was applied to breast and prostate cancer, two of the most common cancers in the United States. For the four breast cancer biomarkers ER, PR, HER2, and Ki-67, the disagreement rate against quality-assured registry values in 1,852 breast cancer cases was lowest for ER at 1.67% and highest for Ki-67 at 9.18%. In 920 prostate cancer cases, the disagreement rate with the deployed models stayed below 8% for every categorical Gleason variable, while the exact-value PSA metric reached 17.28%. PSA therefore generated the largest volume of candidate discrepancies, though individual PSA flags require more corroboration than categorical flags. Applied to a full year of records (11,740 breast and 6,792 prostate cases), the framework flagged disagreements and assigned each a probable cause, grounded in NAACCR coding rules, to prioritize expert review. Extracted values are linked to their supporting text, and model, prompt, and pipeline versions are logged for reproducibility.

By directing review toward records most likely to contain discrepancies, the framework offers an efficient, auditable alternative to random QA sampling for cancer registries.

## 1 Introduction

Cancer registries are a critical resource for cancer surveillance, epidemiologic research, quality improvement, and outcomes research ^1^. In the United States, population-based cancer surveillance is supported primarily by the CDC’s National Program of Cancer Registries (NPCR) ^2^ and the National Cancer Institute’s Surveillance, Epidemiology, and End Results (SEER) Program ^3^, while the Commission on Cancer’s National Cancer Database (NCDB) ^4^ provides a complementary hospital-based source of oncology data. Together, these programs provide broad population coverage, and the U.S. Cancer Statistics database now includes more than 42 million cases diagnosed since 2001^5^ and more than 1.6 million new diagnoses each year ^1^. These data support incidence and survival estimates, studies of treatment patterns and comparative effectiveness, and, increasingly, biomarker-informed research ^6,7^. Their scientific and public health value, however, depends on the accuracy, completeness, consistency, and timeliness of the information they contain ^8,9^.

Maintaining high-quality registry data is challenging because many important variables must be manually abstracted from unstructured clinical documents, including pathology reports, operative notes, and laboratory records ^8,10^. Standardized coding rules and registrar training help improve consistency across facilities and registries ^2,4^, but quality assurance (QA) still relies heavily on human review by Oncology Data Specialists (ODS). Cancer surveillance in the United States operates at two levels ^2^. Hospital-based registries abstract and code data on patients diagnosed or treated at a single facility, while central (population-based) registries aggregate these submissions across all reporting facilities in a defined geographic area to produce complete, standardized population-level data. Central registries are responsible for consolidating and quality-assuring the data they receive, and it is here that ODS review hospital submissions against coding standards and source documentation. Registrars must read narrative reports, identify relevant clinical information, assign structured values, and verify that registry entries agree with the source documentation ^11^. This process is labor-intensive or time-consuming and remains subject to human error and variation between reviewers ^9,12^.

Registry QA is further limited by the small proportion of records that can feasibly be re-reviewed. Although established quality-control frameworks emphasize completeness, validity, and reliability ^9,11,13^, routine expert review typically covers fewer than 10% of abstracted records and often relies on random sampling ^13^. Random review is not well suited to identifying systematic errors. When a limited review effort is spread across the entire registry, relatively few records from any one facility, time period, or registrar may be examined. Recurring coding problems can therefore remain undetected, while substantial reviewer effort is spent confirming records that are already correct. Errors in biomarker coding, missing information, or inconsistent scales and units may then persist and affect downstream analyses ^9,12^. As the volume and complexity of clinical data continue to grow, a more targeted approach to registry QA is needed.

Many studies have explored methods for converting unstructured clinical text into structured cancer information. Early studies combined clinical natural language processing (NLP) with knowledge representation to extract disease characteristics from pathology reports ^10^, and later systems extended these approaches to cancer phenotyping and clinical-concept extraction from oncology narratives ^14^. More recently, large language models (LLMs) have been applied to a wide range of oncology extraction tasks ^15^, including cancer staging ^16^, extraction of clinical and survival-related information from pathology reports ^17^, fine-grained staging descriptors ^18^, temporal relations in oncology records ^19^, and structured findings from breast cancer pathology and ultrasound reports ^20^. LLMs have also been used to populate discrete variables for cancer database construction ^21^, and comparative studies suggest that they can match or outperform traditional NLP approaches for extracting complex information, including biomarkers, from free text ^22^. Together, these studies show that LLMs can reliably extract many clinically relevant variables from oncology documentation.

Beyond extraction alone, systems have been built to help registrars code more efficiently, ranging from hybrid NLP and rule-based tools that suggest a code for the registrar to confirm ^23^, to fully automated classifiers that assign codes directly and route only uncertain cases for review ^24^. These tools, however, are designed to assist coding at the time of abstraction, and they are evaluated by how accurately they reproduce codes against a reference standard. The question facing cancer registries is different: their biomarker variables have already been abstracted by trained registrars, and the practical need is to decide which completed records most warrant re-review. To our knowledge, LLM extraction has not been evaluated at population-registry scale as an independent audit of required biomarker fields, with discrepancies used to direct expert review.

In this study, we develop an evaluation-driven LLM framework for cancer registry quality assurance. We evaluate the approach in breast and prostate cancer, two of the most commonly diagnosed cancers in the United States ^25^, focusing on clinically important biomarkers routinely collected by cancer registries. The selected biomarkers span a range of documentation standardization, from consistently recorded categorical results to more variably documented continuous values, making them a practical setting for developing and evaluating an automated QA approach before extending it to more variable clinical information. Section 2 describes the framework’s architecture and the data, models, and statistical methods used to evaluate it. Section 3 reports extraction performance against quality-assured registry values for both cancers, applies the framework at population scale to surface candidate discrepancies, and introduces a downstream classification step that assigns a probable cause to each flagged case. Section 4 discusses the implications of these findings for registry quality assurance, and Section 5 concludes with the framework’s limitations and directions for future work.

## 2 Methods

### 2.1 Data Source and Study Cohorts

This study draws data from the Missouri Cancer Registry, a central registry that aggregates submissions from more than 100 reporting facilities. For each case, the registry holds both structured coded fields and a set of unstructured free-text fields. These free-text fields are not the original clinical documents; they are dense, abstracted summaries entered by the reporting facility’s cancer registrar, condensing the pathology report, laboratory results, diagnostic procedure notes, staging workup, and clinician remarks into the registry’s submission format. Because this abstraction step can omit details present in the original documents, it places an upper bound on what any downstream extraction method automated or manual can recover from the registry submission alone.

Given this constraint, we designed the evaluation around two cohorts that differ in whether their registry values have already undergone quality assurance:

- *Ground-truth validation cohort*. A subset of records already quality-checked and validated by the expert review, a certified ODS responsible for registry coding and quality assurance, whose registry values served as the reference standard for model evaluation. This cohort comprised 1,852 breast cancer cases and 920 prostate cancer cases.
- *Registry-scale cohort*. The full 2023 diagnosis-year cohort for each site, comprising 11,740 breast cancer cases and 6,792 prostate cancer cases, for which registry values have not undergone this quality-assurance review. This cohort is used to demonstrate the framework’s operation at population scale and to surface candidate discrepancies for expert re-review.

#### Breast cancer biomarkers

Estrogen receptor (ER) and progesterone receptor (PR) status indicate whether a tumor’s growth is hormone-driven and determine eligibility for endocrine therapy. Human epidermal growth factor receptor 2 (HER2) status identifies tumors eligible for HER2-targeted therapy. The Ki-67 proliferation index is a continuous percentage reflecting the proportion of proliferating tumor cells, used as a marker of tumor growth rate.

#### Prostate cancer biomarkers

Gleason score and its component patterns describe tumor differentiation on a pathologist-assigned ordinal scale, reported separately from biopsy (clinical) and prostatectomy or autopsy (pathological) specimens, with an optional tertiary pattern recorded when a third, less common growth pattern is present. Prostate-specific antigen (PSA) is a continuous laboratory value used for diagnosis, risk stratification, and monitoring.

### 2.2 Framework Architecture

The auditing framework proceeds through a set of connected layers (Figure 1). A data layer combines the unstructured clinical documentation described above with the structured registry fields, which serve as the reference standard against which extracted values are later compared. An LLM extraction layer then pulls clinically relevant information from the unstructured text (Section 2.3). Once extracted, both the LLM outputs and the raw registry codes pass through a normalization layer that maps them onto a shared representation for comparison; normalization rules are specific to each variable (Section 2.4).

**Figure 1.**
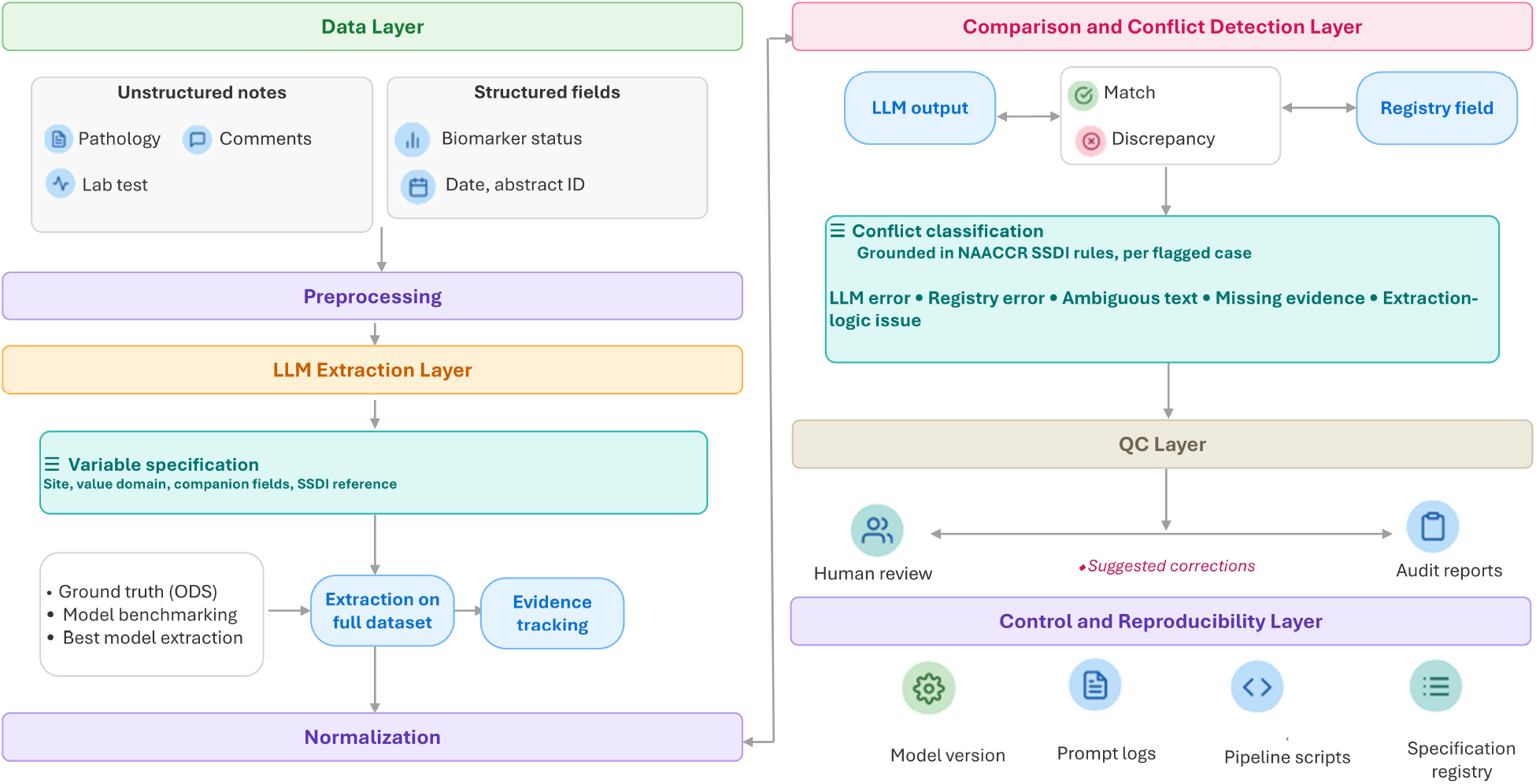
An LLM-based framework for quality assurance in cancer registries.

With both sources normalized, a comparison and conflict-detection layer classifies each case by agreement between the two sources, flagging discordant cases for review while cases of agreement or mutual unavailability require no further action.

Flagged discrepancies move to a quality-control layer built around human review (Section 2.5). Underlying the entire process, a control and reproducibility layer keeps a record of model versions, prompt configurations, normalization rules, and pipeline scripts, so that any result can be traced back to the exact pipeline configuration that produced it.

### 2.3 LLM-Based Extraction

With the shared representation defined above, the extraction layer is responsible for producing, from unstructured clinical text, the values that this representation compares against the registry.

#### 2.3.1 Extraction Prompts and Model Selection

Prompts were tailored to each cancer type and biomarker. Each prompt specifies a closed set of allowed output codes and the rules governing their extraction, aligned with NAACCR coding standards where relevant. For each case, the full clinical text used for extraction is retained alongside the predicted values, so that any flagged case can be reviewed against its source documentation without re-reading the complete record.

Since the registry variables could be directly identified in the source text, we used a zero-shot prompting strategy for extraction. For more challenging variables with greater variation in how values were reported, we also piloted few-shot prompting using a small, cost-efficient model. Specifically, we tested five to six examples for PSA, which showed the greatest textual variability, using Mistral Small-24B. Few-shot prompting did not improve accuracy compared with zero-shot prompting. On inspection, the model also appeared to follow the wording of the provided examples too closely rather than generalizing across the diverse phrasing in the source text. We therefore used zero-shot prompting for the final extraction.

Candidate models were first evaluated on the manually reviewed ground-truth dataset. The models were compared based on overall accuracy and consistency across biomarkers. The model with the highest overall accuracy and the most balanced performance across biomarkers was then selected for application to the corresponding registry-scale cohort.

#### 2.3.2 Models and Computing Environment

To meet institutional privacy and security requirements, we tested only open-source LLMs that can be deployed locally in a HIPAA-compliant environment: MedGemma-27B, Mistral Small-24B, Llama 3.3-70B, Gemma 4-31B, Gemma 4-12B, and Gemma 4-E4B. Models were served locally through the LM Studio API using a zero-shot, prompt-based extraction strategy without task-specific fine-tuning. Model outputs were returned in structured JSON; when biomarker information was absent or indeterminate, the value was coded as “Unknown” per the predefined extraction schema. All experiments ran on a local workstation with two NVIDIA RTX A6000 GPUs (48 GB VRAM each), an AMD Ryzen Threadripper PRO 5955WX processor (16 cores), and 128 GB of system memory. All cancer sites were evaluated with the same models and hardware.

#### 2.3.3 Statistical Analysis

For each biomarker, every case was assigned to one of five mutually exclusive categories: agreement on the same known value (Known-Known Same, *a*), a conflict between two known values (Known-Known Different, *b*), a value present in the registry but not returned by the model (Registry-only, *c*), a value returned by the model but absent from the registry (LLM-only, *d*), and cases where neither source returned a value (Unknown-Unknown, *e*), with *a* + *b* + *c* + *d* + *e* = *N*. Overall accuracy is the proportion of cases in agreement, counting both matching known values and jointly-unknown Overall accuracy cases (Equation 1), and the disagreement (flag) rate is its complement (Equation 2):

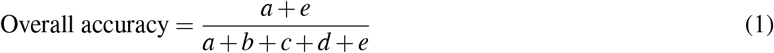

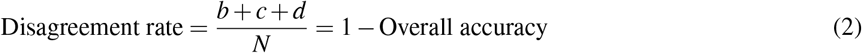

For binary biomarkers (ER, PR, HER2), precision, recall, and F1-score are also reported on the known-known subset (the *a* + *b* cases), using the registry value as the reference standard. These measures are not defined for the ordinal Gleason variables or the continuous PSA value (Section 3.2.1).

Accuracy proportions are reported with 95% Wilson confidence intervals (Equation 3), where 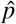 is the observed accuracy and *z* = 1.96:

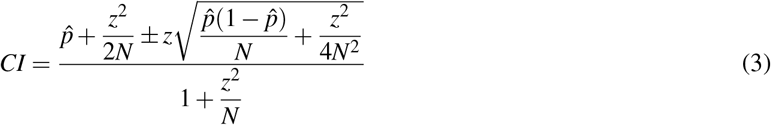

For the ordinal Gleason variables we additionally computed quadratic-weighted Cohen’s *κ* (*κ*_*w*_) over the known-known cases only (the *a* + *b* cases where both sources returned a value), across the ordered categories of the variable. Disagreements are penalized by the square of their distance on the ordinal scale (Equation 4), so that adjacent-grade near-misses count far less than distant ones:

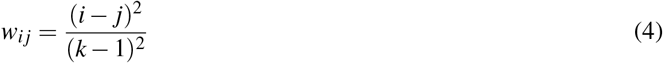

where *i* and *j* index the *k* ordered categories. The weighted *κ* is computed from the observed (*O*) and chance-expected (*E*) known-known counts (Equation 5):

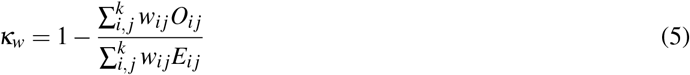

Overall accuracy (Equation 1) is computed over all *N* cases and therefore counts registry-only and LLM-only cases as disagreements, whereas *κ*_*w*_ (Equation 5) is computed only over the *a* + *b* known-known cases and does not reflect abstention. A *κ*_*w*_ near 1.0 alongside a lower overall accuracy therefore indicates near-perfect agreement on committed values combined with many registry-only cases (values the model left unknown), not a discrepancy between the two metrics. Because *κ*_*w*_ requires a fixed set of ordered categories, it is not reported for the continuous PSA measurement.

*κ*_*w*_ is reported as a complementary measure of agreement quality and is not used for model selection, which is based on overall accuracy alone. For the ordinal Gleason variables, we do not report macro-averaged precision, recall, or F1: these categories are numerous and highly imbalanced, with several grades represented by only one or two cases in the ground-truth cohort, so macro-averaged measures are dominated by near-empty classes and misrepresent performance that overall accuracy and *κ*_*w*_ capture more faithfully.

### 2.4 Normalization

Because the LLM’s extracted values and the registry’s raw codes are represented differently, both are mapped onto a shared representation before comparison. All applicable NAACCR Site-Specific Data Item (SSDI) coding rules were followed for each biomarker; given the length and detail of the SSDI Manual, only the rules most relevant to comparison are summarized below. Normalization rules are specific to each biomarker.

#### ER and PR

Registry: 1 (positive), 0 (negative), 7/9 (unknown). LLM output: POS, NEG, Unknown. Both are mapped to the same three-way representation.

#### HER2

Immunohistochemistry (IHC) score 0 or 1+ → negative; 3+ → positive; 2+ (equivocal) → resolved using a reported POS/NEG status if available, otherwise negative.

Handling biomarker values also required accounting for variations in how they appeared in the source text. For PSA, numeric values were sometimes concatenated with nearby dates, so a preprocessing step was used to separate the PSA value from the date before extraction. For Ki-67, the source text sometimes reported a range (e.g., “70-80%”) rather than a single value. These ranges were kept as reported rather than converted to a midpoint, and a case was considered a match when the registry’s single value fell within the reported range.

#### Ki-67

Retained as a continuous percentage. Registry values are single numbers; LLM output may be a single value or a range (e.g., “70-80%”), which is preserved rather than averaged. A case is concordant if the registry value falls within the reported range.

#### Gleason score and patterns

The exact pattern and score were extracted according to all applicable NAACCR coding rules, including the source-priority rule specifying that, when both biopsy and prostatectomy/autopsy values are available, the prostatectomy/autopsy value is authoritative.

#### PSA

A single numeric value in the range 0-1000 ng/mL was extracted exactly as reported.

### 2.5 Quality Control and Conflict Diagnosis

For each flagged case, the system generates an audit report that summarizes the conflict, surfaces the relevant source text extracted earlier, and proposes a correction for the reviewer to accept or revise. To help prioritize this review, we also designed a downstream conflict-diagnosis step that assigns each flagged case a probable cause, grounded in the relevant NAACCR Site-Specific Data Item rules: LLM error, registry error, ambiguous source text, missing evidence, or an extraction-logic issue. This component is described further in the Results.

### 2.6 Reproducibility

The control and reproducibility layer keeps a versioned record of every run, so any result in this paper can be traced back to the exact model, prompt, and pipeline configuration that produced it. This record also underlies the audit reports generated for each flagged case: when an ODS reviews a discrepancy, the same versioned configuration that produced the flagged value is available for inspection, so a reviewer can confirm not only what the model predicted but under what conditions. Because this record is structured and versioned, it could also support a dashboard summarizing flag volumes, review status, and model performance across variables and cancer sites over time, helping registries monitor the framework’s output without inspecting individual audit reports one by one.

## 3 Results

### 3.1 Breast Cancer Biomarkers

Breast cancer was selected as the initial use case because it represents the most frequently reported cancer type in the Missouri Cancer Registry, providing a large and diverse cohort for evaluation. The extraction task focused on four key breast cancer biomarker variables required by the NAACCR: ER, PR, HER2, and Ki-67 proliferation index, normalized as described in Section 2.4.

#### 3.1.1 Ground-Truth Validation Cohort (N = 1,852)

The ground-truth validation cohort for breast cancer comprised 1,852 cases.

Table 1 summarizes zero-shot biomarker extraction accuracy across all six evaluated LLMs on the ground-truth validation cohort. Mean overall accuracy exceeded 94% for every model, ranging from 94.63% to 95.97%. Among them, Gemma 4-31B consistently achieved the highest or near-highest accuracy on every biomarker (ER, PR, HER2, and Ki-67), in addition to the highest mean overall accuracy. Based on these results, Gemma 4-31B was selected for deployment in the registry-scale auditing phase for the breast cancer biomarkers.

**Table 1.** Zero-shot biomarker extraction accuracy (%) across six open-source large language models on the breast cancer ground-truth validation cohort (N = 1,852).

| Model | ER (%) | PR (%) | HER2 (%) | Ki-67 (%) | Mean overall accuracy |
| --- | --- | --- | --- | --- | --- |
| MedGemma-27B | 97.79 | 96.22 | 95.52 | 89.90 | 94.86 |
| Mistral Small-24B | 97.68 | 95.79 | 95.57 | 89.74 | 94.70 |
| Llama 3.3-70B | 98.54 | 97.62 | 96.17 | 90.55 | 95.72 |
| Gemma 4-31B | 98.33 | 97.73 | 96.98 | 90.82 | 95.97 |
| Gemma 4-12B | 97.30 | 95.79 | 95.95 | 90.28 | 94.83 |
| Gemma 4-E4B | 97.52 | 96.00 | 96.11 | 88.88 | 94.63 |

Detailed extraction performance metrics for the selected model are presented in Table 2. Overall accuracy was 98.33% for ER, 97.73% for PR, and 96.98% for HER2, and lower for Ki-67 (90.82%), reflecting a higher proportion of cases in which one or both sources lacked a definitive value.

**Table 2.** Breast cancer biomarker extraction performance, Gemma 4-31B, on the ground-truth validation cohort (N = 1,852). Confidence intervals computed using Wilson’s method.

| Biomarker | N Total | N Known-Only | Accuracy (Known-Only) | Overall Accuracy | 95% CI | Precision | Recall | F1 Score |
| --- | --- | --- | --- | --- | --- | --- | --- | --- |
| ER | 1,852 | 1,759 | 99.43 | 98.33 | 0.9763-0.9882 | 99.80 | 99.53 | 99.66 |
| PR | 1,852 | 1,715 | 99.13 | 97.73 | 0.9695-0.9832 | 99.38 | 99.46 | 99.42 |
| HER2 | 1,852 | 1,488 | 99.13 | 96.98 | 0.9609-0.9766 | 96.57 | 96.02 | 96.30 |
| Ki-67 | 1,852 | 774 | 95.35 | 90.82 | 0.8942-0.9205 |  |  |  |

Table 3 decomposes disagreement cases by conflict category. For ER and PR, direct Known-Known conflicts were rare (10 and 15 cases, respectively), indicating high concordance when both sources provided a value. The predominant category of disagreement for HER2 was Registry Known / LLM Unknown (23 cases). For Ki-67, the Registry Unknown / LLM Known category was largest (71 cases), reflecting cases where the LLM extracted a value absent from the registry.

**Table 3.** Disagreement category breakdown: Gemma 4-31B on the breast cancer ground-truth validation cohort (N = 1,852).

| Biomarker | KK Same | KK Different | Registry Known / LLM Unknown | Registry Unknown / LLM Known | Unknown-Unknown | Total Disagree | Disagree Rate (%) |
| --- | --- | --- | --- | --- | --- | --- | --- |
| ER | 1,749 | 10 | 16 | 5 | 72 | 31 | 1.67 |
| PR | 1,700 | 15 | 20 | 7 | 110 | 42 | 2.27 |
| HER2 | 1,475 | 13 | 23 | 20 | 321 | 56 | 3.02 |
| Ki-67 | 738 | 36 | 63 | 71 | 944 | 170 | 9.18 |

#### 3.1.2 Registry-Scale Validation (N = 11,740)

The framework was then applied to the complete 2023 calendar year of breast cancer records (N = 11,740), the most recent completed year in the registry, which had not undergone quality assurance.

Gemma 4-31B was subsequently applied to the full cohort of 11,740 breast cancer cases (Table 4). Overall disagreement rates were relatively low across all four biomarkers, consistent with the high concordance observed in the ground-truth validation. Disagreement rates were 4.21% for ER, 4.93% for PR, 5.47% for HER2, and 10.57% for Ki-67, mirroring the relative ordering seen in the validation cohort.

**Table 4.** Registry-scale discrepancy analysis, Gemma 4-31B, applied to the full breast cancer cohort (N = 11,740).

| Biomarker | Total Cases | Disagreements (N) | Disagreement Rate (%) |
| --- | --- | --- | --- |
| ER | 11,740 | 494 | 4.21 |
| PR | 11,740 | 579 | 4.93 |
| HER2 | 11,740 | 642 | 5.47 |
| Ki-67 | 11,740 | 1,241 | 10.57 |

To better characterize disagreement patterns, registry-scale results were categorized according to the five-class comparison scheme (Figure 2). Across all four biomarkers, complete agreement (Known-Known, Same) represented most cases, demonstrating strong consistency between information in unstructured clinical documentation and structured registry records. Direct Known-Known conflicts (value mismatches) were relatively uncommon across all biomarkers. A larger proportion of discordant cases involved asymmetric availability, in which one source reported a value and the other reported it as unknown (registry-only or LLM-only, Figure 2).

**Figure 2.**
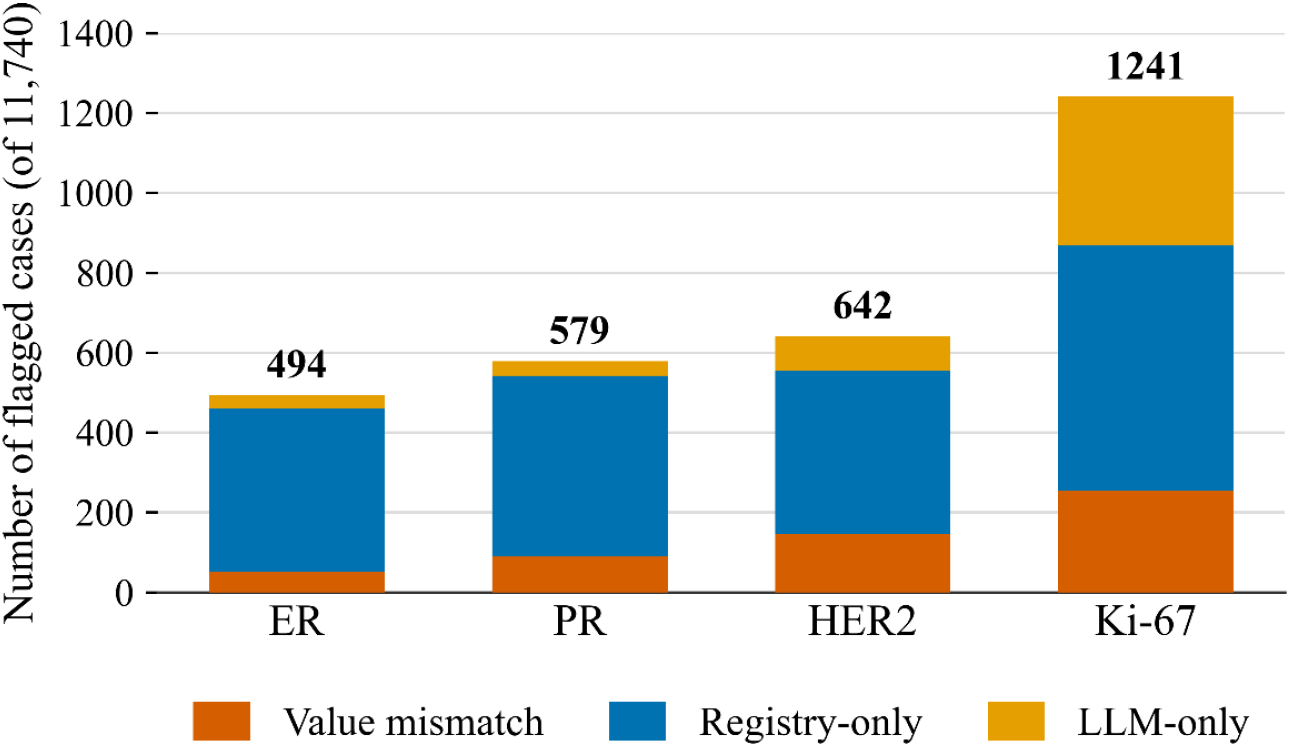
Disagreement category distributions by biomarker across the full breast cancer cohort (N = 11,740). Bars show, per biomarker, the number of flagged cases partitioned into value mismatches, registry-only extractions, and LLM-only extractions.

#### 3.1.3 Triple-Negative Status as a Composite Endpoint

Beyond single biomarkers, we evaluated the framework on triple-negative breast cancer (TNBC) status, a composite endpoint defined as ER-negative, PR-negative, and HER2-negative. TNBC is clinically consequential because it is ineligible for endocrine and anti-HER2 therapies and carries a distinct prognosis ^26^, so a miscoded receptor that changes triple-negative status has direct treatment implications. A case was classified as triple-negative only when all three receptors were a confirmed negative; cases in which HER2 was equivocal or unrecorded were treated as indeterminate and not counted as triple-negative.

Applied to the full 2023 breast cancer cohort (N = 11,740), the framework and the registry showed high concordance on triple-negative status: 1,177 cases (10.03%) were triple-negative in the registry and 1,129 (9.62%) by the model, with the two sources agreeing on 1,090 cases. They diverged on 126 cases (1.07% of the cohort), of which 87 were triple-negative in the registry but not the model and 39 by the model but not the registry. This bidirectional pattern shows that the framework surfaces both candidate registry omissions and its own extraction limitations, rather than errors in only one direction.

These discordances arose from two distinct mechanisms, a distinction that matters for how they are triaged. In 48 of the 87 registry-only cases, the model returned no value for one or more receptors rather than a conflicting value, indicating limited extraction coverage rather than a contradiction with the registry. The remaining discordances reflected genuine value conflicts, in which one source recorded a positive receptor that would exclude the triple-negative classification asserted by the other; for example, in 9 cases the model extracted a positive HER2 result where the registry had recorded the case as triple-negative, and in 9 cases the registry recorded a positive ER. Because the registry-scale cohort has not undergone quality-assurance review, these cases carry no reference standard and are not adjudicated automatically; instead, because a change in triple-negative status carries direct therapeutic consequences, all 126 discordant cases were routed to an ODS for targeted re-review, with cases involving a conflicting positive receptor prioritized above those arising from missing extractions.

This analysis illustrates that the framework surfaces clinically consequential discrepancies cases where an apparent miscoding would change triple-negative status at a scale and specificity that random-sample QA is unlikely to achieve: 126 targeted cases drawn from a cohort of 11,740, concentrated on the determination most likely to affect treatment eligibility.

### 3.2 Prostate Cancer

#### 3.2.1 Ground-Truth Validation Cohort (N = 920)

A ground-truth validation cohort of 920 prostate cancer cases was assembled from the Missouri Cancer Registry using the same methodology as for breast cancer (Section 2.1); ODS-validated registry values served as the reference standard.

All six models were evaluated on the prostate-specific registry variables: Gleason score and Gleason patterns from both clinical (biopsy) and pathological (prostatectomy/autopsy) assessments, Gleason tertiary pattern, and PSA laboratory value. Since a single case’s clinical text fields may contain information from both a needle biopsy and a subsequent radical prostatectomy, the extraction prompts incorporated the applicable NAACCR source-priority rules for each variable. For variables defined as clinical, information from the clinical workup was used, whereas variables defined as pathological were extracted according to their specified pathological source requirements. For prostate cancer variables such as pathological Gleason Score and Gleason Patterns, values from the radical prostatectomy or autopsy were prioritized over earlier biopsy findings.

Tables 5 and 6 report overall accuracy and *κ*_*w*_ for the clinical (biopsy) and pathological (prostatectomy/autopsy) variables, respectively; Figures 3 and 4 decompose the disagreements into value mismatches, registry-only extractions, and LLM-only extractions.

**Table 5.** Overall accuracy (%) and quadratic-weighted *κ* (*κ*_*w*_) on clinical variables across six open-source LLMs (N = 920). Highest value in each column is shown in bold. PSA *κ*_*w*_ is omitted (see note).

| Model | Gleason Score |  | Gleason Patterns |  | PSA Lab Value |
| --- | --- | --- | --- | --- | --- |
| | Acc. | $\kappa_w$ | Acc. | $\kappa_w$ | Acc. |
| Gemma 4-31B | **95.11** | 0.961 | **93.37** | **0.982** | **82.72** |
| Gemma 4-12B | 94.13 | 0.950 | 91.41 | 0.967 | 82.61 |
| Gemma 4-E4B | 90.33 | 0.919 | 90.22 | 0.964 | 80.00 |
| Llama 3.3-70B | 93.91 | **0.972** | 91.52 | 0.975 | 69.24 |
| MedGemma-27B | 91.30 | 0.955 | 81.74 | 0.954 | 81.85 |
| Mistral Small-24B | 93.15 | 0.969 | 91.85 | 0.966 | 81.20 |

Overall accuracy = agreement (identical known values, or unknown on both sides) ÷ 920 reports. $\kappa_w$ = quadratic-weighted Cohen’s $\kappa$ over reports where both sources reported a value. $\kappa_w$ is not reported for PSA because it is a continuous measurement.

**Table 6.** Overall accuracy (%) and quadratic-weighted *κ* (*κ*_*w*_) on pathological variables across six open-source LLMs (N = 920). Highest value in each column is shown in bold.

| Model | Gleason Score |  | Gleason Patterns |  | Tertiary Pattern |  |
| --- | --- | --- | --- | --- | --- | --- |
| | Acc. | $\kappa_w$ | Acc. | $\kappa_w$ | Acc. | $\kappa_w$ |
| Gemma 4-31B | 89.67 | **1.000** | 89.24 | 0.989 | 96.30 | **1.000** |
| Gemma 4-12B | 89.35 | **1.000** | 88.80 | 0.987 | 95.65 | **1.000** |
| Gemma 4-E4B | 88.70 | **1.000** | 88.37 | **0.990** | 94.89 | 0.900 |
| Llama 3.3-70B | 93.15 | 0.991 | 92.07 | 0.966 | 96.30 | 0.879 |
| MedGemma-27B | **94.35** | 0.969 | **92.39** | 0.942 | 97.50 | **1.000** |
| Mistral Small-24B | 90.98 | 0.907 | 90.43 | 0.917 | **97.61** | 0.637 |

Overall accuracy = agreement ÷ 920 reports. $\kappa_w$ computed over reports where both sources reported a value.

**Figure 3.**
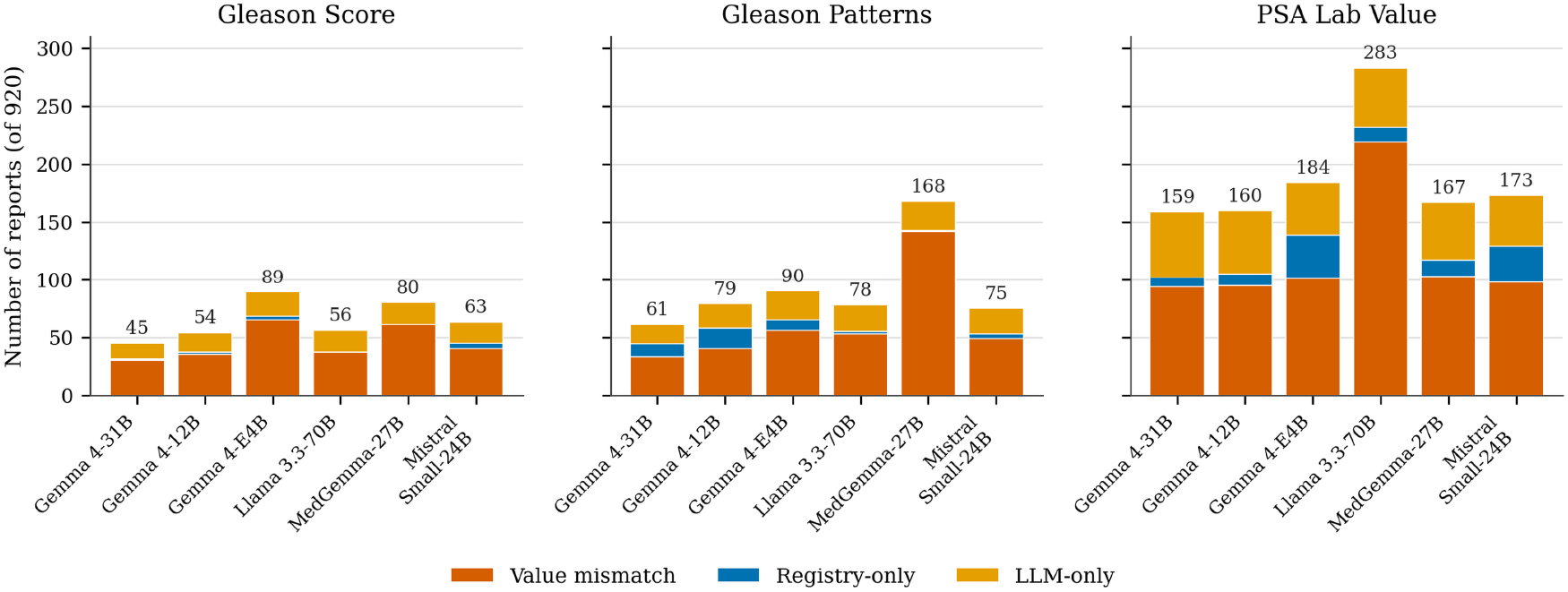
Composition of disagreements on clinical variables across six open-source LLMs (N = 920), for the two Gleason variables and the PSA laboratory value.

**Figure 4.**
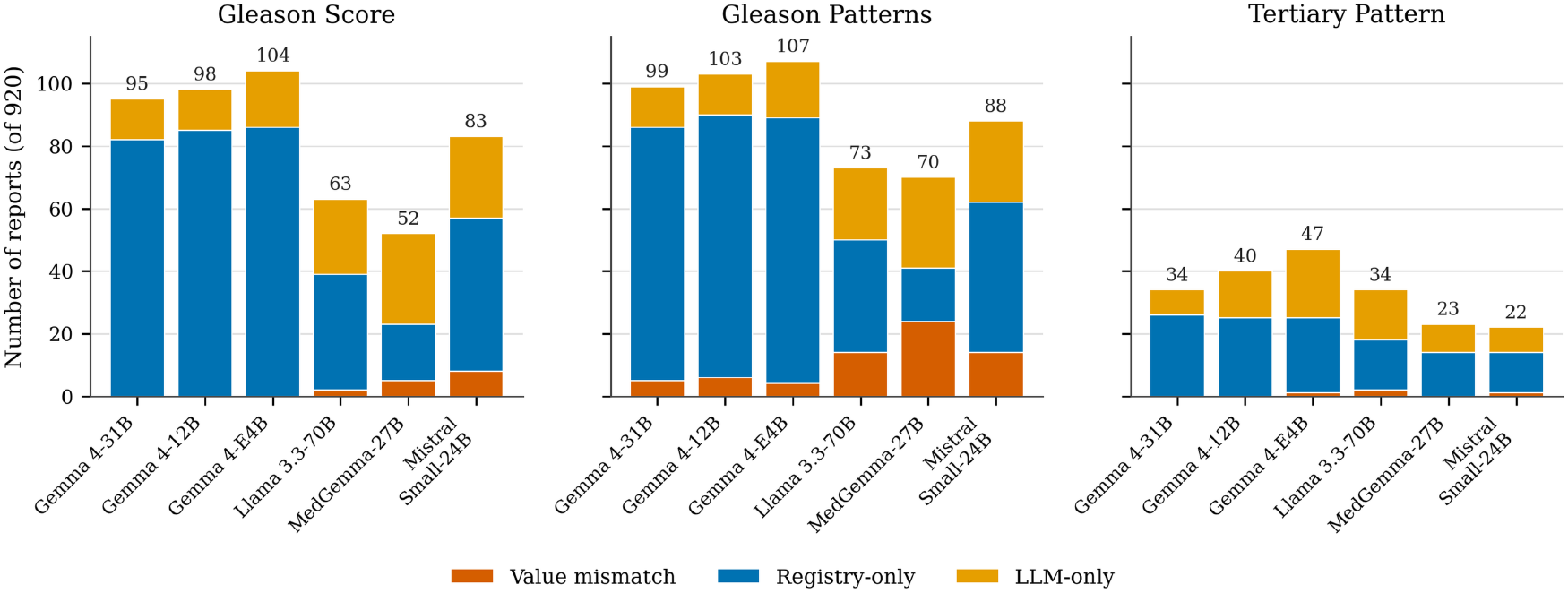
Composition of disagreements on pathological variables across six open-source LLMs (N = 920).

##### Clinical variables

On clinical variables (Table 5), Gemma 4-31B achieved the highest overall accuracy on both Gleason score (95.11%) and Gleason patterns (93.37%), followed closely by several other models. Weighted agreement was uniformly high: *κ*_*w*_ exceeded 0.95 for every model on Gleason patterns and 0.91 on Gleason score, placing all six in the “almost perfect” range. MedGemma-27B’s exact-match accuracy on Gleason patterns was comparatively low (81.74%), while its *κ*_*w*_ remained high (0.954).

Exact-value agreement on the PSA laboratory value was substantially lower than on the Gleason variables, as anticipated for a continuous measurement transcribed across heterogeneous note formats. Overall exact-match accuracy ranged from 69.24% (Llama 3.3-70B) to 82.72% (Gemma 4-31B), and the disagreements were dominated by value mismatches rather than registry-only or LLM-only extractions (Figure 3). Llama 3.3-70B was a clear outlier, with 219 value mismatches, more than double any other model, indicating a systematic difference in how it transcribed or rounded numeric PSA values. Because PSA is continuous, quadratic-weighted *κ* is not reported; PSA generated the largest volume of candidate discrepancies of any prostate variable.

##### Pathological variables

On pathological variables (Table 6), MedGemma-27B achieved the highest overall accuracy on Gleason score (94.35%) and patterns (92.39%), while the Gemma 4 models, despite lower overall accuracy, reached near-perfect weighted agreement once they committed to a value (*κ*_*w*_ = 1.000 on Gleason score). Their conflicts were dominated by registry-only cases (81-86 per variable for Gleason score and patterns, and 24-26 for the tertiary pattern) rather than value mismatches (0-6 per variable; Figure 4), consistent with lower accuracy driven mainly by abstention rather than incorrect extraction. No single model was best on every report type and biomarker, so model selection for registry-scale deployment was made per report type: Gemma 4-31B for the clinical (biopsy) variables and MedGemma-27B for the pathological (prostatectomy/autopsy) variables. This configuration was carried forward to the registry-scale prostate auditing phase.

#### 3.2.2 Registry-Scale Validation

The two selected models were applied to a full year of prostate cancer cases (N = 6,792 unique patients) that had not undergone quality-assurance review. For each variable, we report the agreement rate (the proportion of cases where the model and registry agree) and the flag rate (the proportion of cases routed for review), along with the composition of the flags.

Agreement between the deployed models and the registry was high and closely tracked the validation-cohort results (Table 7), supporting the stability of the framework at scale. For the categorical Gleason variables, agreement ranged from 90.28% to 97.53% and flag rates from 2.47% to 9.72%, meaning that between roughly one in forty and one in ten cases were surfaced for review. The PSA laboratory value again behaved differently: exact-value agreement was 80.30%, producing 1,338 flags (19.70% of cases), by far the largest review burden of any variable, consistent with the difficulty of exact-value transcription observed during validation.

**Table 7.** Registry-scale agreement and discrepancy summary for the one-year prostate cohort (N = 6,792 unique patients), by deployed model and variable.

| Report type | Variable | Model | Agreement (95% CI) | Flagged n | Flag rate | % mismatch |
| --- | --- | --- | --- | --- | --- | --- |
| Clinical | Gleason Score | Gemma 4-31B | 92.24 (91.58-92.85) | 527 | 7.76% | 25.4 |
|  | Gleason Patterns | Gemma 4-31B | 90.36 (89.63-91.04) | 655 | 9.64% | 29.5 |
|  | PSA (exact value) | Gemma 4-31B | 80.30 (79.34-81.23) | 1,338 | 19.70% | 51.5 |
| Pathological | Gleason Score | MedGemma-27B | 92.17 (91.50-92.78) | 532 | 7.83% | 9.0 |
|  | Gleason Patterns | MedGemma-27B | 90.28 (89.56-90.96) | 660 | 9.72% | 27.3 |
|  | Tertiary Pattern | MedGemma-27B | 97.53 (97.13-97.87) | 168 | 2.47% | 3.6 |

Agreement = proportion of cases on which the model and the registry concur (matching known values or jointly unknown). Flagged n = cases routed for ODS review (any disagreement). Flag rate = flagged n ÷ 6,792. % mismatch = value mismatches ÷ flagged n. PSA is evaluated on exact-value matching.

The composition of the flags (Figure 5) is directly relevant to the review workload, because the three categories differ in how likely they are to represent a genuine registry error. Value mismatches, where both the registry and the model report a value but the two differ, are the highest-yield flags; registry-only and LLM-only extractions, where only one source reports a value, are weaker signals that more often reflect extraction coverage than registry error. The proportion of flags that were value mismatches varied markedly by variable: it was highest for PSA (51.5%) and lowest for the categorical pathological variables (3.6-9.0% for Gleason score and the tertiary pattern), where most flags were asymmetric-availability cases. This indicates that the review yield is concentrated in the PSA and Gleason-pattern flags, and that a substantial share of the categorical-variable flags can be triaged as lower priority.

**Figure 5.**
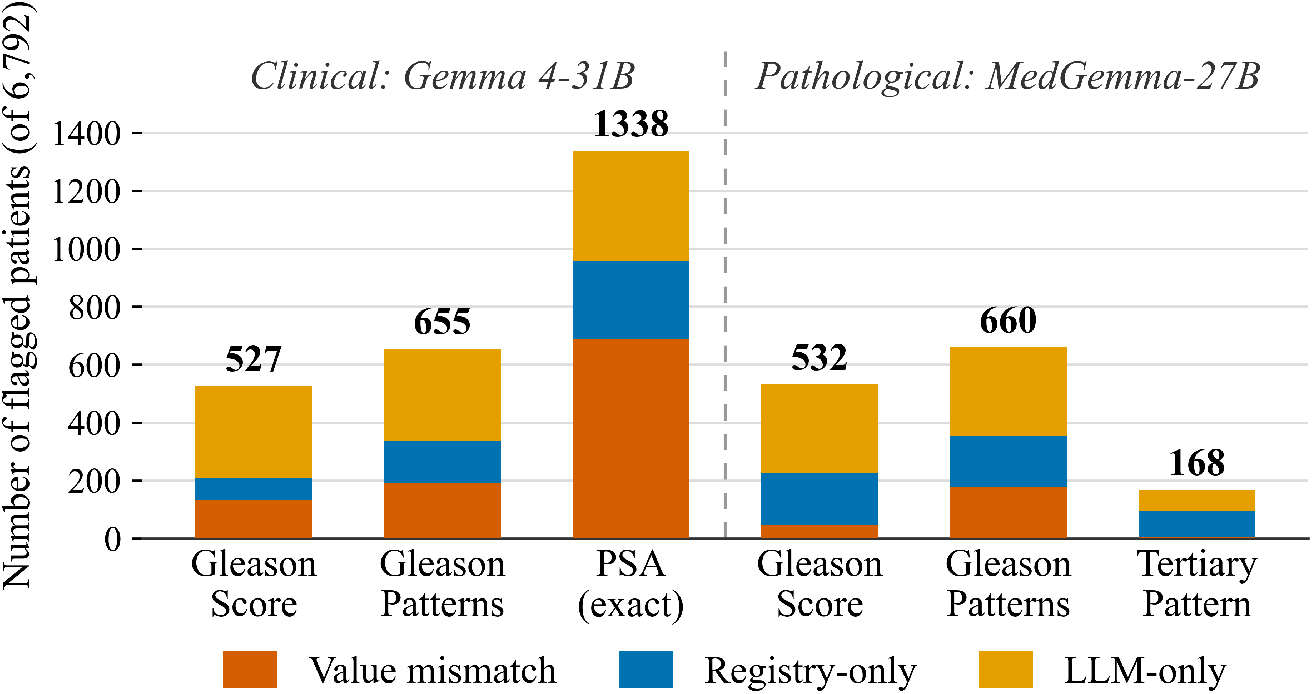
Composition of flagged discrepancies in the one-year registry-scale run (N = 6,792) for the deployed configuration (Gemma 4-31B on clinical variables, MedGemma-27B on pathological variables).

### 3.3 Automated Conflict Diagnosis

The registry-scale run identifies discrepancies but does not yet explain them; each flagged case is currently routed to an ODS for manual adjudication. As an early pilot, prior to the design of this component, we conducted a blinded human review of a random sample of flagged ER/PR conflicts using Mistral Small-24B, a model whose ground-truth accuracy on these biomarkers (Table 1) was within 1-2 percentage points of the deployed Gemma 4-31B. An Oncology Data Specialist, given only the abstract ID, the registry value, and the model’s extracted value, judged which source was correct without knowledge of either value’s origin. Of 77 reviewed conflicts, the registry was judged correct in 30 (39%), the model in 22 (29%), the available documentation was itself too ambiguous for the reviewer to determine a correct value in 23 (30%), and in 2 cases (3%) neither the registry nor the model value was judged correct. This pilot motivated the design of a downstream Case Diagnosis Layer (Figure 6) that assigns a probable cause to each flagged case. It is implemented as a single-shot LLM classifier: for each conflict it assembles a fixed context (the source report text, the registry value, and the model’s extracted value) and returns one of five structured categories (LLM extraction error, registry coding error, ambiguous source text, insufficient evidence, or extraction-logic issue). Because the assembly and output schema are fixed and each case requires a single model call, the component is fully auditable, which suits its intended use across multiple cancer types and variables.

**Figure 6.**
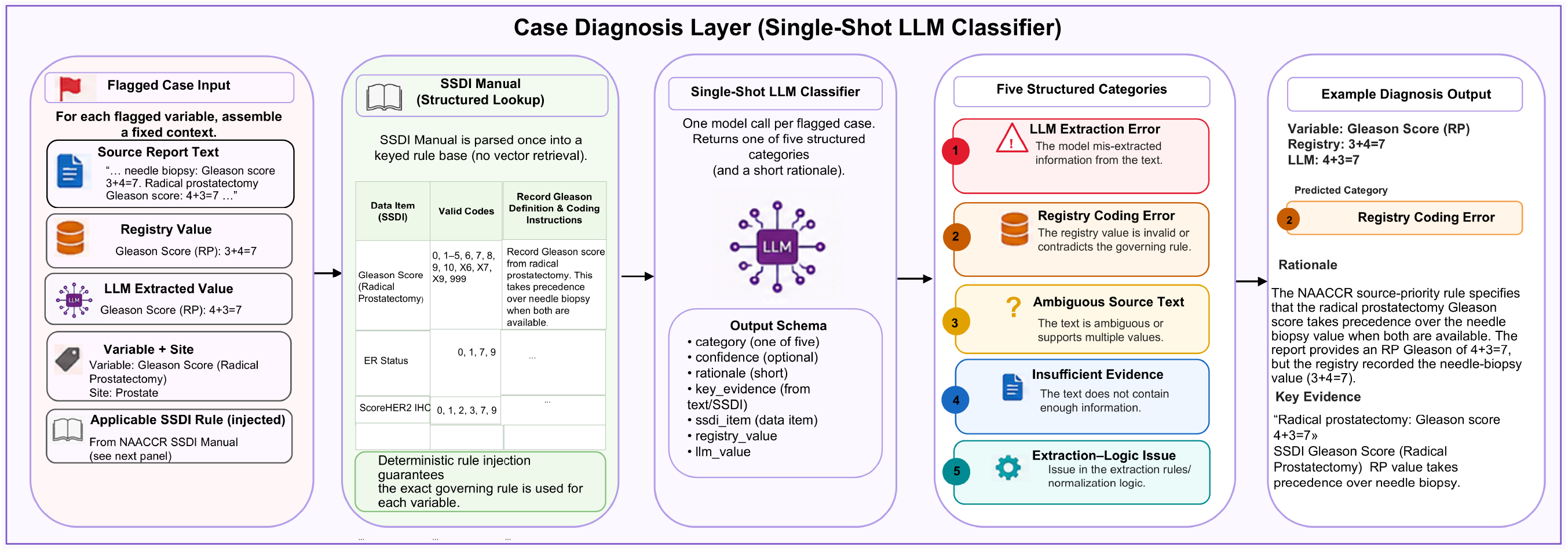
Design of the Case Diagnosis Layer. Each flagged conflict is combined with a fixed context and the matching NAACCR SSDI coding rules, parsed once into a keyed lookup, then classified by a single-shot LLM into one of five structured categories. The example is illustrative (a schematic case, not a real record).

This layer is meant to do more than simply label individual cases. By assigning a likely cause to each flagged discrepancy, we can look across a variable and understand why it behaves the way it does. For example, a high registry-only rate might be caused by gaps in extraction, unclear documentation, or a consistent coding practice. A single agreement measure would not show these differences. Separating the causes also helps prioritize the discrepancies that are most likely to be actual registry errors for expert review, rather than spending limited ODS effort on issues caused by extraction problems or unavoidable ambiguity. In this sense, the diagnosis layer serves as the final step in a triage process: starting with all records, narrowing them down to flagged discrepancies, then to value mismatches, and finally to the cases most likely to reflect a registry error.

To ground the classification in registry coding rules, each flagged variable is linked to its corresponding NAACCR Site-Specific Data Item (SSDI). Rather than vector-based retrieval, the SSDI Manual ^7^ is parsed once into a keyed lookup (valid codes, definition, and coding instructions per data item), and the relevant rules are injected deterministically into the classification context. This structured-lookup design retrieves the exact governing rule for each variable and never surfaces an incorrect section (important for compliance-sensitive coding), and it directly strengthens the categories: a registry value that is not a valid code for its SSDI is strong evidence of a coding error, and likewise for the model’s output. Source-priority conflicts such as a case in which the registry appears to have coded a needle-biopsy Gleason value where the NAACCR rule specifies that the radical-prostatectomy value should take precedence, are one clinically important class of case this layer is designed to surface.

As a preliminary demonstration, we applied the Case Diagnosis Layer to a stratified random sample of 200 value-mismatch conflicts (the registry-known / model-known discordant category) drawn across all breast and prostate variables, using Gemma 4-31B with the full NAACCR SSDI coding rules injected as described above. The classifier assigned a registry coding error in 131 cases (65.5%), an LLM extraction error in 54 (27%), an extraction-logic issue in 11 (5.5%), ambiguous source text in 3 (1.5%), and insufficient evidence in 1 (0.5%). The predominance of registry-coding-error assignments over extraction-error assignments is consistent with the layer’s intended purpose of surfacing genuine registry discrepancies rather than merely re-flagging model mistakes, while the substantial minority attributed to the model indicates that the classifier does not attribute every conflict to the registry.

These category assignments are the model’s own judgments and are not yet independently validated; the sample was drawn only from value-mismatch conflicts, so the distribution characterizes that subset rather than all flagged discrepancies.

## 4 Discussion

### Principal Findings

In this study, we developed and evaluated an LLM-based auditing framework for automated quality assurance of cancer registry biomarker data in a population-based cancer registry. Across both breast and prostate cancer biomarkers, zero-shot extraction from unstructured clinical text showed strong agreement with registry-validated values, exceeding 95% known-only accuracy for all four breast biomarkers and staying below an 8% disagreement rate with the deployed models for every categorical Gleason variable in the ground-truth cohort, indicating that large language models can serve as a reliable independent source of evidence for registry quality assurance. In addition to identifying individual biomarker discrepancies, the framework also detected a small number of clinically consequential discrepancies involving triple-negative breast cancer classification and Gleason source-priority coding, which may be missed by routine random-sample review. Together, these results support the framework’s central premise: that targeted, evidence-linked auditing could help direct limited expert review toward records that warrant further attention, rather than distributing review uniformly across the registry.

### Model Selection and Performance

The registry text contains protected patient information. All models were therefore run locally rather than through cloud services, limiting our evaluation to open-weight models that could run entirely inside the institution. We deliberately evaluated models spanning different sizes and architectures (compact models, larger general-purpose models, and reasoning-oriented models) rather than a single system, to assess how much model size influences performance on this task. This speaks to a practical question for registries: does reliable biomarker auditing need a large model, or can a smaller one that is cheaper to run do just as well? In our results, larger model size did not consistently translate into better performance: the largest model evaluated (Llama 3.3-70B, 70B parameters) was never the top performer in overall accuracy on any prostate variable (Tables 5 and 6), and was a clear outlier for PSA transcription (Section 3.2.1). Its only leading result was on breast ER (Table 1: 98.54% vs. 98.33% for the runner-up, Gemma 4-31B), a margin of 0.21 percentage points. Across the remaining nine biomarker/variable comparisons, the best-performing model was consistently mid-sized Gemma 4-31B (31B parameters), MedGemma-27B (27B parameters), or Mistral Small-24B (24B parameters), suggesting that a lighter, less demanding model may be sufficient for this kind of auditing, which matters when hardware is limited. The runtimes and throughput reported here are specific to this hardware configuration and would differ on other systems, depending on the available GPU, memory, and concurrency. This would not change the extracted values themselves: for the non-reasoning models, extraction is deterministic regardless of hardware.

### Interpreting Disagreement: Ambiguity, Abstention, and Source Priority

Not every discrepancy the framework surfaces has a clear answer. This is not always a limitation of the model. In some cases, the underlying text is genuinely ambiguous or poorly formatted, making it difficult for even a human reader to confidently determine the correct value. A stray dash before “ER” in the registry’s abstracted text is one example: it may mean the biomarker is negative, or it may just be a formatting artifact with no clinical meaning. The text alone cannot settle it, and this is not an isolated problem: in an early pilot review of flagged ER/PR conflicts (Section 3.3), the reviewer judged the documentation too ambiguous to determine a correct value in 30% of cases. Messy notes, inconsistent formatting, and incomplete documentation all produce disagreements that reflect the state of the source record rather than a failure of extraction. These cases are therefore not errors to correct but records to escalate: the system’s role is to route them for expert review rather than force a decision. Seen this way, they are a useful output in their own right, pointing to exactly the records where the documentation is too unreliable for any automated method to be trusted.

Gleason reporting also showed a clear difference between clinical and pathological variables. Agreement was somewhat lower on pathological variables (Tables 5 and 6), but the disagreement breakdown shows why: most of the gap came from cases where the model returned “unknown,” not from cases where the model and registry gave different known values. For pathological Gleason score, MedGemma-27B (the model deployed for pathological variables) had only five known-known conflicts against eighteen cases where the registry had a value and the model did not. These two situations call for different follow-up: a value the model could not commit to points to unclear documentation or limited extraction, while two conflicting known values points to a possible coding error and warrants expert review. This distinction matters beyond bookkeeping. A model that answers “unknown” when the text is unclear may reduce unnecessary flags and keep expert review focused on cases with stronger evidence of a conflict; a model that always commits may achieve higher coverage but risk introducing additional erroneous values. This is why the framework reports accuracy and *κ*_*w*_ together, and separates every disagreement into value mismatches versus one-sided abstentions, instead of reducing model behavior to one accuracy number.

PSA showed the weakest exact-value agreement of any prostate variable (Table 5), and unlike the Gleason variables, most PSA disagreements were value mismatches rather than one-sided abstentions. This fits PSA’s nature: it is a continuous value reported in inconsistent formats across notes, which makes exact-value extraction harder than extracting a category. PSA therefore produced the largest volume of candidate discrepancies of any prostate variable, but that same difficulty means individual PSA flags carry more uncertainty than categorical flags and should be corroborated, for example, by the Case Diagnosis Layer or expert review. Llama 3.3-70B stood out with far more value mismatches than any other model, despite having more than twice the parameter count of the best-performing models. This fits a broader pattern: parameter count alone does not determine LLM performance. Newer architectures, training data, and training methods let smaller models match or beat older, larger ones, a trend termed “capability density,” shown to roughly double every three months among open-source models ^27^. Llama 3.3 predates the newer Gemma 4 and MedGemma architectures used here, which may be one possible explanation for its weaker PSA extraction despite its size. A related point applies to the source-priority conflicts the framework can surface: a flagged case should not be assumed to be a coding error, since the available documentation may not include everything the original registry decision was based on.

### Generalizability and Future Extension

Finally, the framework’s architecture supports expansion without redesigning it. Because each cancer–biomarker combination is defined by its own prompt and normalization rule and dispatched through a shared lookup, adding NAACCR biomarkers for new cancer sites, or additional categorical variables like the Gleason variables evaluated here, extends the same pipeline rather than requiring a new one. As a result, the same unmodified pipeline produced both the breast cancer results (four biomarkers) and the prostate cancer results (six biomarkers) reported above, simply by adding new prompts and normalization rules rather than new code, a design that extends in the same way to biomarkers for other cancer sites. The Case Diagnosis Layer follows the same pattern: because it retrieves coding rules from a keyed NAACCR lookup rather than a variable-specific implementation, extending diagnosis to a new biomarker requires only that the biomarker’s SSDI entry exist in the lookup, not new classification logic. This modularity is what allowed a single evaluation cycle to cover ten biomarkers across two cancer sites, and is the basis for the multi-cancer, continuous-QA system we envision as future work.

Future work will extend the framework to additional cancer sites and registry variables. For explicitly stated biomarkers, zero-shot extraction was already sufficient, and the added cost of fine-tuning is unlikely to be justified given the framework’s goal of targeted error detection rather than maximal extraction accuracy. Fine-tuning may instead be warranted for variables that require clinical interpretation rather than direct transcription, where zero-shot performance is more likely to be limited.

## 5 Conclusion

This study presents an auditable LLM-based framework for quality assurance of cancer registry biomarker data in a population-based cancer registry. The framework shifts quality assurance from undirected random sampling toward targeted, risk-based validation by prioritizing records with potential inconsistencies between registry data and clinical documentation.

Using a zero-shot, prompt-based approach with locally run open-source LLMs, the system produced a low disagreement rate against ODS-validated biomarker values: a known-only rate at or below 5% across all four breast cancer biomarkers and at or below 1% for ER, PR, and HER2, and an overall rate below 8% for the deployed models’ prostate Gleason variables, while identifying clinically relevant discrepancies at registry scale. Rather than replacing human registrars, the framework functions as a decision-support tool that directs expert review to the cases most likely to contain meaningful errors.

A key feature of the framework is its full auditability: retained source text, model configurations, and processing logs ensure transparency and reproducibility, enabling reliable traceability of extracted values to source clinical text. The results demonstrate that LLM-based auditing can reduce manual workload while maintaining human oversight, and that it can surface clinically consequential discrepancies such as those affecting triple-negative breast cancer classification that are statistically unlikely to be detected by conventional 10% random-sample review.

Future work will extend the framework to additional cancer sites and registry variables, and will explore fine-tuning, retrieval-augmented generation, and uncertainty estimation to further improve extraction reliability. Ultimately, we envision a scalable, multi-agent system enabling continuous, transparent, and risk-adaptive quality assurance across population-based cancer registries.

## Declarations

### Author Contributions

Maryam Seifaddini (Conceptualization, Study design, Implementation, Writing original draft), Mohammad Beheshti (Study design, Implementation, Writing review and editing), Iris Zachary (Study design, Writing review and editing, Supervision, Resources), Jeffrey Steffens (Ground-truth preparation, Validation), Deborah Carnagey (Ground-truth preparation, Validation), Magda Esebua (Pathology expertise, Clinical review, Writing review and editing), and Mark Wakefield (Urology expertise, Clinical review, Writing review and editing).

### Data Availability

The data underlying this study contain protected health information (PHI) from the Missouri Cancer Registry and cannot be made publicly available. Qualified researchers may request access to de-identified data through the Missouri Cancer Registry, subject to a data use agreement.

## Supplementary Material

**Table S1.** Exact composition of disagreements underlying Figures 3 and 4, prostate cancer ground-truth cohort (N = 920), all six open-source LLMs.

| Report / Biomarker | Model | Mismatch | Registry-only | LLM-only | Total |
| --- | --- | --- | --- | --- | --- |
| Clinical Gleason Score | Gemma 4-31B | 30 | 1 | 14 | 45 |
|  | Gemma 4-12B | 35 | 2 | 17 | 54 |
|  | Gemma 4-E4B | 65 | 3 | 21 | 89 |
|  | Llama 3.3-70B | 37 | 0 | 19 | 56 |
|  | MedGemma-27B | 61 | 0 | 19 | 80 |
|  | Mistral Small-24B | 40 | 5 | 18 | 63 |
| Clinical Gleason Patterns | Gemma 4-31B | 33 | 11 | 17 | 61 |
|  | Gemma 4-12B | 40 | 18 | 21 | 79 |
|  | Gemma 4-E4B | 56 | 9 | 25 | 90 |
|  | Llama 3.3-70B | 53 | 2 | 23 | 78 |
|  | MedGemma-27B | 142 | 1 | 25 | 168 |
|  | Mistral Small-24B | 49 | 4 | 22 | 75 |
| Clinical PSA Lab Value | Gemma 4-31B | 94 | 8 | 57 | 159 |
|  | Gemma 4-12B | 95 | 10 | 55 | 160 |
|  | Gemma 4-E4B | 101 | 38 | 45 | 184 |
|  | Llama 3.3-70B | 219 | 13 | 51 | 283 |
|  | MedGemma-27B | 103 | 14 | 50 | 167 |
|  | Mistral Small-24B | 98 | 31 | 44 | 173 |
| Pathological Gleason Score | Gemma 4-31B | 0 | 82 | 13 | 95 |
|  | Gemma 4-12B | 0 | 85 | 13 | 98 |
|  | Gemma 4-E4B | 0 | 86 | 18 | 104 |
|  | Llama 3.3-70B | 2 | 37 | 24 | 63 |
|  | MedGemma-27B | 5 | 18 | 29 | 52 |
|  | Mistral Small-24B | 8 | 49 | 26 | 83 |
| Pathological Gleason Patterns | Gemma 4-31B | 5 | 81 | 13 | 99 |
|  | Gemma 4-12B | 6 | 84 | 13 | 103 |
|  | Gemma 4-E4B | 4 | 85 | 18 | 107 |
|  | Llama 3.3-70B | 14 | 36 | 23 | 73 |
|  | MedGemma-27B | 24 | 17 | 29 | 70 |
|  | Mistral Small-24B | 14 | 48 | 26 | 88 |
| Pathological Tertiary Pattern | Gemma 4-31B | 0 | 26 | 8 | 34 |
|  | Gemma 4-12B | 0 | 25 | 15 | 40 |
|  | Gemma 4-E4B | 1 | 24 | 22 | 47 |
|  | Llama 3.3-70B | 2 | 16 | 16 | 34 |
|  | MedGemma-27B | 0 | 14 | 9 | 23 |
|  | Mistral Small-24B | 1 | 13 | 8 | 22 |

## References

[1] M. C. White, F. Babcock, N. S. Hayes, A. B. Mariotto, F. L. Wong, B. A. Kohler, and H. K. Weir. The history and use of cancer registry data by public health cancer control programs in the United States. Cancer, 123(S24): 4969–4976, 2017.

[2] Centers for Disease Control and Prevention. National Program of Cancer Registries (NPCR). https://www.cdc.gov/national-program-cancer-registries/index.html, 2026. Accessed Sept 8, 2026.

[3] National Cancer Institute. Surveillance, Epidemiology, and End Results (SEER) program overview. https://seer.cancer.gov/about/overview.html, 2026. Accessed Sept 8, 2026.

[4] B. E. Palis, L. M. Janczewski, A. E. Browner, J. Cotler, L. Nogueira, L. C. Richardson, V. Benard, R. J. Wilson, N. Walker, R. M. McCabe, D. J. Boffa, and H. Nelson. The National Cancer Database conforms to the standardized framework for registry and data quality. Ann Surg Oncol, 31:5546–5559, 2024.

[5] Centers for Disease Control and Prevention. United States Cancer Statistics (USCS) public use database: about the data. https://www.cdc.gov/united-states-cancer-statistics/public-use/about.html, 2026. Accessed Sept 8, 2026.

[6] B. Aryal, Z. Bizhanova, E. A. Joseph, Y. Yin, P. L. Wagner, E. Dalton, W. A. LaFramboise, D. L. Bartlett, and C. J. Allen. Navigating precision oncology: insights from an integrated clinical data and biobank repository initiative across a network cancer program. Cancers, 16(4):760, 2024.

[7] North American Association of Central Cancer Registries. Site-specific data item (ssdi) manual, version 3.3. https://www.naaccr.org/wp-content/uploads/2025/12/SSDI-Manual-v3.3_printed.pdf, 2025. Accessed Sept 14, 2026.

[8] O. M. Jensen, D. M. Parkin, R. MacLennan, C. S. Muir, and R. G. Skeet. Cancer Registration: Principles and Methods. IARC Scientific Publications No. 95. International Agency for Research on Cancer, Lyon, 1991.

[9] D. M. Parkin and F. Bray. Evaluation of data quality in the cancer registry: principles and methods. Part II: completeness. Eur J Cancer, 45:756–764, 2009.

[10] A. Coden, G. Savova, I. Sominsky, et al. Automatically extracting cancer disease characteristics from pathology reports into a disease knowledge representation model. J Biomed Inform, 42:937–949, 2009.

[11] D. M. Parkin and F. Bray. Evaluation of data quality in the cancer registry: principles and methods. Part I: comparability, validity and timeliness. Eur J Cancer, 45:747–755, 2009.

[12] A. Caldarella, G. Amunni, C. Angiolini, et al. Feasibility of evaluating quality cancer care using registry data and electronic health records: a population-based study. Int J Qual Health Care, 24:411–418, 2012.

[13] F. Bray, A. Znaor, P. Cueva, et al. Quality control at the population-based cancer registry. In Planning and Developing Population-Based Cancer Registration in Low-Income and Middle-Income Settings, chapter 5. International Agency for Research on Cancer, Lyon, 2014. IARC Technical Report No. 43.

[14] T. Duong and T. Thieu. OncoNLP: cancer comprehend annotation, a pipeline for cancer phenotype and clinical extraction. In 2024 IEEE EMBS International Conference on Biomedical and Health Informatics (BHI), pages 1–6, Piscataway, NJ, 2024. IEEE.

[15] M. Seifaddini, M. Beheshti, S. Richberg, M. Esebua, and I. Zachary. From information extraction to clinical reasoning: a systematic scoping review of large language models in cancer pathology reports. Mod Pathol, 39(9): 101043, 2026.

[16] C. H. Chang, M. M. Lucas, G. Lu-Yao, and C. C. Yang. Classifying cancer stage with open-source clinical large language models. In 2024 IEEE 12th International Conference on Healthcare Informatics (ICHI), pages 76–82, Piscataway, NJ, 2024. IEEE.

[17] Y. C. Chang, S. H. Hsiao, W. C. Yeh, Y. C. Hsing, C. C. Wang, and C. Y. Chen. Extracting critical clinical indicators and survival prediction of lung cancer from pathology reports using large language models. Comput Biol Med, 195: 110621, 2025.

[18] H. Cho, S. Yoo, B. Kim, et al. Extracting lung cancer staging descriptors from pathology reports: a generative language model approach. J Biomed Inform, 157:104720, 2024.

[19] V. Sharma, A. Fernandez, A. Ioanovici, D. Talby, and F. Buijs. Lexicans at ChemoTimelines 2024: leveraging large language models for temporal relations extraction in oncological electronic health records. In Proceedings of the 6th Clinical Natural Language Processing Workshop, pages 394–405, 2024.

[20] H. S. Choi, J. Y. Song, K. H. Shin, J. H. Chang, and B. S. Jang. Developing prompts from large language model for extracting clinical information from pathology and ultrasound reports in breast cancer. Radiat Oncol J, 41:209–216, 2023.

[21] M. Gilbert, A. Crutchfield, and J. Luo. Using a large language model for automated extraction of discrete elements from clinical notes for creation of cancer databases. Int J Radiat Oncol Biol Phys, 120(2S):e625, 2024.

[22] J. Joseph-Thomas, P. Keswarpu, N. Singh, K. Jiwani, B. A P. Gurha, S. Tiwari, A. Ashwani, V. Samal, V. Agarwal, M. Lawrence, and R. Martin. Comparing traditional NLP methods and LLM-based extraction for identifying biomarkers in lung cancer. J Clin Oncol, 43(16_suppl):e13607, 2025.

[23] H.-J. Dai, C.-C. Chen, T. H. Mir, et al. Integrating predictive coding and a user-centric interface for enhanced auditing and quality in cancer registry data. Comput Struct Biotechnol J, 24:322–333, 2024.

[24] L. Gondara, J. Simkin, S. Devji, G. Arbour, and R. Ng. ELM: a hybrid ensemble of language models for automated tumor group classification in population-based cancer registries. arXiv:2503.21800, 2025.

[25] R. L. Siegel, T. B. Kratzer, N. S. Wagle, H. Sung, and A. Jemal. Cancer statistics, 2026. CA Cancer J Clin, 76(1): e70043, 2026.

[26] W. D. Foulkes, I. E. Smith, and J. S. Reis-Filho. Triple-negative breast cancer. N Engl J Med, 363(20):1938–1948, 2010.

[27] C. Xiao, J. Cai, W. Zhao, B. Lin, G. Zeng, J. Zhou, Z. Zheng, X. Han, Z. Liu, and M. Sun. Densing law of LLMs. Nat Mach Intell, 7(11):1823–1833, 2025.

